# Efficacy of a Gamified Digital Platform for Substance Use Education and Overdose Prevention Among College Students: a Pilot and Feasibility Study

**DOI:** 10.64898/2026.06.15.26355709

**Authors:** Madeline E. Hilliard, Ryan Foreman, Tahmid Khan, Elizabeth Zona, Ayati Mishra, Sean J. Howse

## Abstract

**Background:** For US young adults aged 18-25 in the 2018-2024 period, fentanyl was involved in 78.2% of the 44,020 unintentional or undetermined-intent overdose deaths, most often co-involving stimulants and other non-opioid substances. While fatal overdose rates in this age group have fallen to their lowest recorded level, emergency medical services-attended non-fatal overdose events have reached record highs, shifting the decisive variable toward bystander recognition and response. College students report near-universal alcohol education but minimal education on the substances actually driving overdose mortality.

**Methods:** We conducted a single-group pre-post evaluation of the DopaGE Portal, a gamified, mastery-based digital platform covering cocaine, MDMA, benzodiazepines, and opioid overdose response, deployed at a public university (UNL) and a multi-campus volunteer network (TACO). Paired pre/post surveys (N=42) measured self-efficacy (7 items; primary), behavioral intentions, risk perception, and knowledge/attitudes on 5-point scales, plus four factual knowledge questions. Paired t-tests, exact McNemar tests, and Benjamini-Hochberg correction across eight primary tests were applied. Institutional naloxone distribution at UNL was tracked as an ecological behavioral outcome. A mandated high-school cohort (N=94) provided supplementary acceptability data.

**Results:** Self-efficacy increased from 2.82 to 4.46 (d=2.00, 95% CI 1.46-2.55; adjusted p<.001), and behavioral intentions from 4.24 to 4.81 (d=1.43; adjusted p<.001), with effects statistically indistinguishable across sites. Three of four knowledge items improved significantly (+31 to +41 percentage points). Risk perception was at ceiling at baseline (4.38/5) and did not change. In the two months following deployment, 38 naloxone kits were distributed on campus (limited to one per person from the campus pharmacy and health center) versus 14 in the preceding two years combined; the campus health center had distributed zero kits in 2025 despite stocked availability. Evaluation ratings were uniformly positive across voluntary and mandated cohorts, with zero negative ratings.

**Conclusions:** A digital-only, gamified intervention produced large gains in overdose-response self-efficacy and substance-specific knowledge, with concurrent campus-level naloxone acquisition consistent with behavioral translation. These findings are preliminary – single-group, modest N, ecological behavioral outcome – and motivate a future randomized controlled trial.

## 1. INTRODUCTION

### 1.1 The losses that persist are fentanyl deaths

Between 2018 and 2024, 595,356 Americans died of unintentional or undetermined-intent drug overdoses; 384,548 of these deaths (64.6%) involved fentanyl (ICD-10 T40.4, synthetic opioids other than methadone – primarily fentanyl and its analogs) [1]. Yet, for young adults ages 18–25 the impact is more devastating: fentanyl was involved in 78.2% of their 44,020 overdose deaths, and its share rose from 61.2% in 2018 to 85.3% in 2023, holding near 80% in 2024 even as total deaths declined [1]. Young-adult overdose mortality has fallen from its 2021 peak (23.5 per 100,000) to its lowest recorded rate (10.8 per 100,000 in 2024) [1]. While the decline is welcome, it is also incomplete as a description of the crisis. The deaths among young adults that remain are overwhelmingly fentanyl deaths; meanwhile the high-risk substance use events that precede death are currently at an all-time high.

### 1.2 The mortality crisis is becoming a survivability crisis

Emergency medical services (EMS)-attended non-fatal overdoses rose in rate per 100,000 from 107.4 in 2018 to 195.7 in the twelve months ending May 24, 2026. This reflects 634,815 non-fatal overdose events in the US between May 25, 2025 and May 24, 2026, now the highest on record [2]. Part of the earlier, pre-pandemic rise reflects expanding jurisdiction participation with the National EMS Information System [3,4], indicating early rates as undercounts and the current record as conservative; with consistently near-complete participation, the rate rose a further 13% from 2023 to current [2]. These two curves, fatal overdose rates declining to a post-pandemic low in 2024 while non-fatal overdoses reached a record high as of May 2026, illuminate the decisive variable at the scene of an overdose itself. Each non-fatal overdose is a survived, life-threatening medical emergency, and the determining factor in whether an event is ultimately fatal or non-fatal increasingly depends on whether a proximate bystander is functionally able and willing to act.

The State Unintentional Drug Overdose Reporting System (SUDORS) data describe typical fatal outcome scenarios for 2024: Two-thirds of decedents (66.6%) had no pulse at the time first responders arrived on scene; only 24.0% had naloxone administered [5]. For 28.6% of overdose deaths, a potential bystander was present and that bystander either took no action or delayed action in supporting the individual whose life was at risk and ultimately lost [5]. When a pulse is already absent at EMS arrival, professional rescue has arrived too late. The national average EMS time to reach the patient is over thirteen minutes for the twelve months ending May 24, 2026, indicating the effective rescuer must already be in the room to prevent a fatality. For college students and young adults, the room is often a residence hall, an apartment, or a party.

### 1.3 Co-involvement calls for caution, contamination variance necessitates

Polysubstance use, concurrent use of more than one drug – regardless of intent [6], is the primary characteristic of recent overdose fatalities: 51.4% of all 2024 overdose deaths involved illegally made fentanyls and one or more other drugs [5]. Among 18-25 year olds whose deaths involved fentanyl between 2018–2024, cocaine was co-involved in 19.9%, psychostimulants – including MDMA, methamphetamine, amphetamines, and methylphenidate – were co-involved in 19.8%, followed by benzodiazepines (15.6%) and alcohol (11.2%) [1]. For all fentanyl-related overdose deaths in the US, cocaine is the drug most frequently co-involved [1,5]. Heroin co-involvement in this age group is comparatively low (8.1%) [1]. Heroin-co-involved deaths largely represent intentional use of an illicit opioid in a supply where fentanyl co-occurs roughly half the time [7], whereas young adults regularly encounter fentanyl through stimulants with no intention of consuming an opioid. Cannabis, the most widely used illicit substance among US young adults [8], appears in just 1.8% of fentanyl-related fatalities [1], suggesting co-involvement tracks pharmacology, not popularity.

On the likelihood that any non-opioid illicit substance will contain fentanyl, there are few validated points of evidence and comparing those available demonstrates no individual can know with certainty. Community drug-checking services report adjusted fentanyl prevalence of 12.5% in powder methamphetamine and 14.8% in powder cocaine, with wide geographic variation [9]; an 11.9-million-sample laboratory analysis found national co-occurrence of ≤1% and ≤4% respectively, yet finds both exceeding 10% in several Northeast states [7]. Product identification by users is no more reliable: among nightclub and festival attendees reporting lifetime ecstasy use who denied ever using novel psychoactive substances, four in ten had hair samples testing positive for such substances. Of the full user group studied, more than one quarter (28.5%) reported discovering that their ecstasy had contained another drug and an additional 21.9% reported suspecting ecstasy they had used had been contaminated [10]. Such contamination is consistent with MDMA and MDA’s collective near-absence (0.2%) from fentanyl-involved deaths, since products sold as ’MDMA’ to the user are categorized in the mortality data as the substances they truly contain [5]. When neither likelihood of contamination nor true substance identity can be known, the protective position is to assume exposure to fentanyl is possible for any illicit substance use, and prepare to respond accordingly.

### 1.4 Access without comprehension is insufficient

The supply-side response to bystander inefficacy in recent years was to make naloxone physically available and legal for anyone to access without a prescription. The FDA approved the first over-the-counter (OTC) nasal naloxone in March 2023 amid extensive media coverage [11]. Yet a nationally representative survey conducted between October and November that same year found that 63.2% of adults ages 18-34 had heard of naloxone, and only 4.7% carried it. The dominant reasons for not carrying naloxone according to those who did not were not knowing where to get it (74.5%) and not knowing how to use it (62.8%) [12]. Prior to naloxone’s OTC designation, state naloxone access laws were similarly not associated with significant reductions in opioid-related overdose deaths among youths (patient-specific laws: IRR 0.97, 95% CI 0.91–1.04) [13]. Name recognition of and legally-supported access to overdose reversal medication, without the knowledge of why and how to obtain and use it, have not produced protected communities. The critical gap is an educational one: awareness and access are insufficient when individuals do not know where to obtain naloxone or how to use it effectively.

### 1.5 Standard university substance use education does not address primary causes of student death

Substance use education for US students is saturated with alcohol content. It sometimes incorporates cannabis, nicotine, or prescription medications, while the remaining substances students may encounter in teenage or early adult years are inconsistently included in such prevention instruction. In pre-intervention surveys across our deployment sites (described below), 94.5% of college students reported prior formal education on alcohol use, while 10.9% reported formal education on naloxone use, 17.3% on cocaine use, 7.3% on benzodiazepine use, and 4.5% education on MDMA use. Meanwhile, the substances and tools most relevant to protecting against current fentanyl mortality trends were the least taught. Incumbent interventions fail along two distinct axes. Directive prohibition messaging, such as the D.A.R.E. model, has been evaluated extensively and does not produce its intended outcomes [14]. Digital programs whose focus surrounds legal substances like alcohol and prescription medications, the AlcoholEdu model, support consistent instruction at scale but avoid addressing illicit substances while utilizing passive instruction: randomized trials show modest knowledge gains and inconsistent behavioral effects that have not been demonstrated beyond 30 days, with substantial attrition concerns [15,16]; current commercial catalogs confirm the model remains alcohol-centric with passive learning architecture [17]. No widely deployed program combines an autonomy-respecting message with an active, mastery-based learning mechanism to address and protect against the illicit substances most likely to produce a fatal outcome; this is the quadrant where developmental science and instructional design could align for the young adult population.

### 1.6 Theoretical basis: manufacturing mastery experience within a developmental window

Self-efficacy theory identifies enactive mastery experience – demonstration of successful performance – as the strongest lever of efficacy beliefs [18]. Mastery learning provides an actionable framework: defined performance thresholds, formative assessment with immediate feedback, corrective exercises, and re-assessment until mastery is demonstrated [19], with meta-analytic support (mean effect size 0.59 across 36 studies [20]) and a motivational architecture, driven by autonomy and competence [19], matched to the central developmental task of ages 18–25 [21,22] where abstract reasoning capacity is still maturing [23]. Digital delivery can match or exceed clinician delivery in adjacent domains [24]. What does not yet exist is rigorous evidence of gamified digital interventions in substance use prevention specifically: a systematic review found mostly methodologically weak studies and called for better-executed evaluation, while maintaining theoretical optimism for gamified health learning despite the lack of evidentiary support found in their review [25]. We understand that review as a caution: testing gamification for this purpose requires methodological precision to justify future controlled trials otherwise lacking in the literature.

### 1.7 The present study

We report a single-group pre-post evaluation of the DopaGE Prevention Portal, a gamified, mastery-based digital platform teaching substance-specific risk education (cocaine, MDMA, benzodiazepines), fentanyl contamination context, alcohol-combination risks, and opioid overdose response including reversal medication administration and rescue breath administration. We evaluate (1) changes in self-efficacy (primary), behavioral intentions, risk perception, and knowledge/attitudes from before to immediately after Portal completion at two sites with distinct motivation profiles; (2) factual knowledge changes on four numeric-recall items; (3) campus-level naloxone acquisition at the primary site as an ecological behavioral outcome; and (4) acceptability across voluntary and mandated cohorts. We hypothesized that students would show large self-efficacy gains even for those with already-high risk perception – such that the Prevention Portal addresses what students lack (actionable capability) regardless of what they already have (awareness of danger).

## 2. METHODS

### 2.1 Study design

Single-group pre-post study with paired surveys at two deployment sites, an ecological institutional-level behavioral outcome at the primary site, and supplementary acceptability data from a third, mandated-participation site. There was no control group; this is acknowledged as a primary limitation and is the explicit motivation for a future randomized controlled trial.

### 2.2 The intervention: DopaGE Prevention Portal

The DopaGE Prevention Portal is a gamified, mastery-based digital platform for substance use education and overdose prevention. Content comprises three substance-focused levels (cocaine, MDMA, benzodiazepines) and an opioid overdose response level covering overdose recognition, naloxone administration, rescue breath administration, and Good Samaritan protections. Each substance-focused level addresses the risk of fentanyl and other contaminants and the risks of simultaneously using the substance with alcohol. Content is animated, scenario-based, and clinically reviewed. Learning follows a mastery structure: students respond to assessment items embedded in and following each level’s interactive video content, receive immediate corrective feedback on every response, must meet defined performance thresholds to progress, and re-encounter previously missed items in a terminal assessment required for completion and certification [19]. The platform is self-paced (approximately 45–60 minutes, completable across one or more sessions) and entirely digital, with no in-person component.

### 2.3 Participants and settings

#### Primary site: University of Nebraska-Lincoln (UNL)

A public research university with Fall 2025 enrollment of 19,378 [26]. The Portal deployed February 1, 2026. First-year students received automatic Portal access; non-first-year students were required to email the student health center to request access, an additional self-selection step. Participation was entirely voluntary. Forty-nine students completed the Portal during the study window (39 first-year, 10 non-first-year); paired pre/post survey data meeting the date criterion were available for 27 (see Section 2.7).

#### Supporting site: TACO

Team Awareness Combating Overdose (TACO) Inc. is an independent 501(c)(3) non-profit organization focused on youth substance use and overdose prevention, operated by undergraduate student volunteers across campuses nationwide, whose primary activity is providing free overdose prevention tools including naloxone. Students complete Portal training as onboarding for volunteer roles (organizational relationships are described in Declarations). Twenty-seven students completed the Portal in the study window; paired data were available for 15.

#### Supplementary site: college-preparatory high school (Dallas–Fort Worth metropolitan area)

A private college-preparatory high school required all 125 second-semester seniors to complete the Portal. The school administered its own surveys, with some similarities to but overall distinct from the DopaGE instrument, and provided those results to DopaGE; 98 students completed pre-surveys (78.4%) and 94 completed post-surveys (75.2%). Because participation was mandated rather than voluntary, this cohort provides an acceptability test free of self-selection; because the instrument differed and responses were collected without individual linkage, its data are reported descriptively and are not included in paired analyses.

### 2.4 Survey instrument

Twenty-eight core items spanning four constructs, plus four factual knowledge questions, evaluation items, and a prior-education inventory, administered at pre (before Portal access) and immediately post (after Portal completion; "iPost"). Scenario-based items at iPost were deliberately reworded from their pre-survey counterparts to reduce recall bias. Ninety-day follow-up is ongoing and is not analyzed here (n=8 complete triplets at the data cutoff).

Construct composites: (1) **Self-efficacy** (7 items; primary outcome): drug avoidance self-efficacy in a party scenario, adapted from the Drug Avoidance Self-Efficacy Scale (DASES) [27]; overdose recognition confidence, adapted from the Opioid Overdose Knowledge Scale (OOKS) [28]; naloxone administration confidence, adapted from the Opioid Overdose Attitudes Scale (OOAS) [28]; cocaine and MDMA safe-decision confidence (DASES-adapted [27]; MDMA risk literature [29]); and substance-specific safety self-assessments for benzodiazepines and MDMA [30,29]. (2) **Risk perception** (6 items), adapted from the PhenX Toolkit Perceived Harm of Substance Use protocol [31,32]. (3) **Behavioral intentions / overdose response readiness** (5 items), adapted from the OOAS [28]. (4) **Knowledge/attitudes** (6 items) [31,27]. All Likert items use 5-point response scales; item phrasing was modified from source instruments for college-student comprehension. (5) **Factual knowledge**: four single-select questions testing recall of specific numeric values taught in the Portal (naloxone duration of action; standard doses of cocaine, alprazolam, and MDMA), developed by DopaGE because no validated instrument covers these facts.

Scoring: Items for which agreement reflects the desired response were scored 1 (Strongly Disagree) to 5 (Strongly Agree); six reverse-keyed items (for which disagreement is desired) are recorded on a negative scale scored −1 (Strongly Disagree) to −5 (Strongly Agree) and were transformed by adding 6 to each value such that all items are reflected by a common 1–5 scale where 5 is the most desirable response. Knowledge questions were scored correct/incorrect.

The pre-Portal inventory asked students to select any of 12 substances/topics on which they had previously received formal education. Evaluation items at iPost assessed perceived real-life usefulness and (from approximately February 21 onward) likelihood of recommending the Portal.

### 2.5 Behavioral outcome: institutional naloxone distribution (UNL)

Naloxone distribution counts were obtained from UNL campus pharmacy records and the student health center. These are institutional-level counts, not linked to individual Portal completers; the comparison is ecological. Historical baseline: 2 kits distributed in calendar year 2024 and 12 in calendar year 2025 (both pharmacy only); the student health center stocked naloxone throughout 2025 and distributed zero kits that year. Four in-person naloxone trainings were held on campus in 2025 (5–50 students per session), did not produce sustained acquisition behavior, and were subsequently discontinued. The health center confirmed that no naloxone-related awareness campaigns, policy changes, or student-facing procedural changes occurred during February–March 2026. Distribution during the study period was limited to one kit per person due to supply constraints.

### 2.6 Anonymity and pairing

Surveys utilized pseudonymous, paired across timepoints by self-generated identification codes. Before each survey, immediately after consent, students answered three personal questions with stable one-word answers (e.g., mother’s first name) and entered the first or last three letters of each, as specifically requested; the nine resulting characters, stored in upper case, form the anonymity tag. Tags are regenerated at every timepoint rather than stored, enabling within-student pairing without collecting any identifying information. No demographic data were collected. Answers or letter choices that vary between surveys allow some imperfect regeneration of non-matching tags, accounting for some Portal completers lacking paired data (Section 3.1).

### 2.7 Analysis populations

Two analysis populations are used, with different denominators. The **education-gap population** comprises all pre-survey respondents at each site within the deployment window (UNL n=81; TACO n=29), used descriptively in the Introduction and for prior-education context; these pulls include students who took the pre-survey regardless of completion status on the Portal education. The **paired population** comprises students with a pre-survey documented on or before March 31, 2026 and a completed iPost survey: 27 at UNL and 15 at TACO (N=42), after excluding one respondent whose response pattern indicated non-genuine data and two students without iPost surveys. All inferential analyses use the paired population.

### 2.8 Ethics

The UNL Institutional Review Board office reviewed the project scope and the Master Services Agreement (MSA) between DopaGE and the institution and determined the project did not meet the threshold requiring full IRB review. The MSA permits collection, use, and analysis of de-identified user data for research, product improvement, and publication of aggregated findings. All data were de-identified; anonymity tags cannot be linked to identities. The applicable exemption basis is 45 CFR 46.104(d)(4) (secondary research using de-identified educational assessment data).

### 2.9 Statistical analysis

Paired t-tests compared pre and iPost composite scores; Cohen’s d for paired data (mean difference divided by the standard deviation of differences) is reported with 95% confidence intervals. Exact McNemar tests compared paired correct/incorrect proportions on the four knowledge questions. The Benjamini-Hochberg procedure controlled false discovery rate across the eight primary tests (four composites, four knowledge questions). Welch’s t-tests compared change scores between sites to assess poolability. Cronbach’s alpha assessed internal consistency of the self-efficacy composite at both timepoints. A pre-specified sensitivity analysis re-estimated the primary outcome on a conservative 4-item composite excluding the ceiling-bound party-scenario item and the two perceived-education items. Naloxone distribution, evaluation items, and prior education are reported descriptively. Primary analyses were conducted in R; an independent verification analysis was run in Python directly against the raw response-level export, reproducing all reported values. Epidemiological statistics cited in the Introduction were generated by the authors from CDC WONDER Multiple Cause of Death queries; full query specifications are provided in Table S4.

## 3. RESULTS

### 3.1 Participant flow and completion characteristics

Of 49 UNL Portal completers, 34 appeared in the survey tracking file, 30 had pre-surveys on or before March 31, and 27 had paired pre/iPost data after exclusions; all 15 TACO pairs fell within the date window (N=42). UNL students predominantly completed pre-survey, Portal, and iPost in a single day (median interval 0 days; 74.1% same-day; mean 2.5 days, range 0–33). TACO students spread completion across days or weeks (median 1 day; 40.0% same-day; mean 6.5 days, range 0–32), consistent with self-paced onboarding into a volunteer role.

**Table 1.** Participant flow and completion characteristics.

|  | UNL | TACO |
| --- | --- | --- |
| Portal completers in window | 49 | 27 |
| Paired pre/iPost (analyzed) | 27 | 15 |
| Median pre-to-iPost interval (days) | 0 | 1 |
| Same-day completion | 74.1% | 40.0% |
| Mean (range) interval, days | 2.5 (0–33) | 6.5 (0–32) |
| Access route | First-years automatic; others by request | Volunteer onboarding |

### 3.2 Construct composites

Self-efficacy internal consistency was good to acceptable (α = 0.822 pre; 0.756 post). Self-efficacy and behavioral intentions improved with very large effects surviving false-discovery correction; risk perception and attitude items, which began near the scale ceiling (4.38 and 4.15 of 5), did not change significantly.

Effects were statistically indistinguishable between sites on every construct (Welch’s tests on change scores, all p>0.26), justifying pooled analysis. Site-specific effects were consistent: self-efficacy d=2.09 at UNL (2.74→4.46) and d=1.82 at TACO (2.98→4.48); behavioral intentions d=1.54 and d=1.25 respectively. Despite markedly different completion pacing (74.1% vs 40.0% same-day), learning outcomes did not differ – the mastery-based structure produced equivalent gains regardless of session pattern.

**Table 2.** Construct composite results (combined sites, N=42).

| Construct | Pre M | Post M | $\Delta$ | Cohen's d [95% CI] | p (raw) | p (BH) |
| --- | --- | --- | --- | --- | --- | --- |
| Self-efficacy (7 items) | 2.82 | 4.46 | +1.64 | 2.00 [1.46, 2.55] | <.0001 | <.001 |
| Behavioral intentions (5 items) | 4.24 | 4.81 | +0.57 | 1.43 [0.99, 1.88] | <.0001 | <.001 |
| Risk perception (6 items) | 4.38 | 4.53 | +0.15 | 0.21 | .17 | .20 |
| Knowledge/attitudes (6 items) | 4.15 | 4.23 | +0.08 | 0.10 | .52 | .52 |

### 3.3 Self-efficacy sensitivity analysis

The 7-item composite includes two items assessing perceived preparedness to help a friend stay safe (benzodiazepine and MDMA safety self-assessments) that showed the largest individual effects (d=2.80 and d=2.01) and could be viewed as manipulation checks; they are retained in the primary composite because they measure confidence to act – a dimension of self-efficacy consistent with Bandura’s framework [18]. A conservative 4-item composite excluding both items and the ceiling-bound party-scenario item yielded d=1.60 (95% CI 1.14–2.05) with internal consistency α=0.816/0.807, confirming that the primary finding is robust to item selection.

**Table 3.** Self-efficacy composite sensitivity analysis (N=42).

| Composite | d [95% CI] | $\alpha$ pre | $\alpha$ post |
| --- | --- | --- | --- |
| 7-item (primary) | 2.00 [1.46, 2.55] | 0.822 | 0.756 |
| 4-item (conservative) | 1.60 [1.14, 2.05] | 0.816 | 0.807 |

### 3.4 Factual knowledge

All four items require recall of specific numeric values (milligram doses; minutes of drug action) – a deliberately demanding test of factual transfer. Three of four improved significantly after false-discovery correction. The cocaine item improved significantly at UNL alone (33.3%→63.0%) but declined slightly at TACO (33.3%→26.7%), yielding combined non-significance.

**Table 4.**
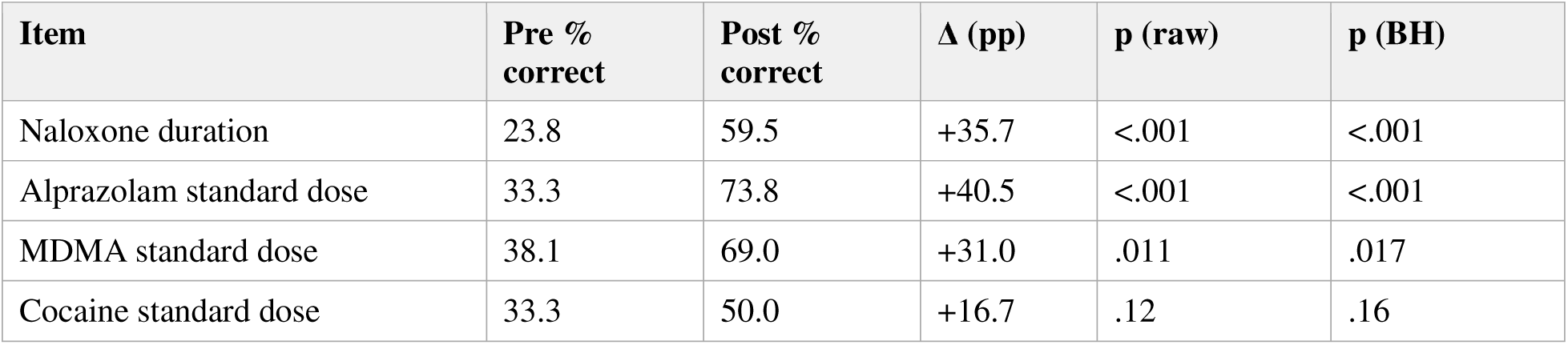
Knowledge question results (combined sites, N=42; exact McNemar tests).

| Item | Pre % correct | Post % correct | $\Delta$ (pp) | p (raw) | p (BH) |
| --- | --- | --- | --- | --- | --- |
| Naloxone duration | 23.8 | 59.5 | +35.7 | <.001 | <.001 |
| Alprazolam standard dose | 33.3 | 73.8 | +40.5 | <.001 | <.001 |
| MDMA standard dose | 38.1 | 69.0 | +31.0 | .011 | .017 |
| Cocaine standard dose | 33.3 | 50.0 | +16.7 | .12 | .16 |

### 3.5 Prior education

Among the paired sample (N=42), 92.9% reported prior formal alcohol education, against 28.6% for opioids, 16.7% for naloxone, 14.3% for cocaine, 9.5% for benzodiazepines, and 4.8% for MDMA. (Figure 1 presents the corresponding distribution among all date-eligible pre-survey respondents, n=110.) The full pre-survey populations show the same structure at larger N: at UNL (n=81), 97.5% alcohol versus 7.4% naloxone and 4.9% MDMA; at TACO (n=29), 86.2% alcohol versus 20.7% naloxone – the elevated naloxone figure consistent with TACO students’ self-selection into an overdose-prevention volunteer organization. The illicit substances most likely to be involved in current overdose mortality are addressed less frequently than non-illicit substances that have little-to-no risk of fentanyl contamination.

**Figure 1.**
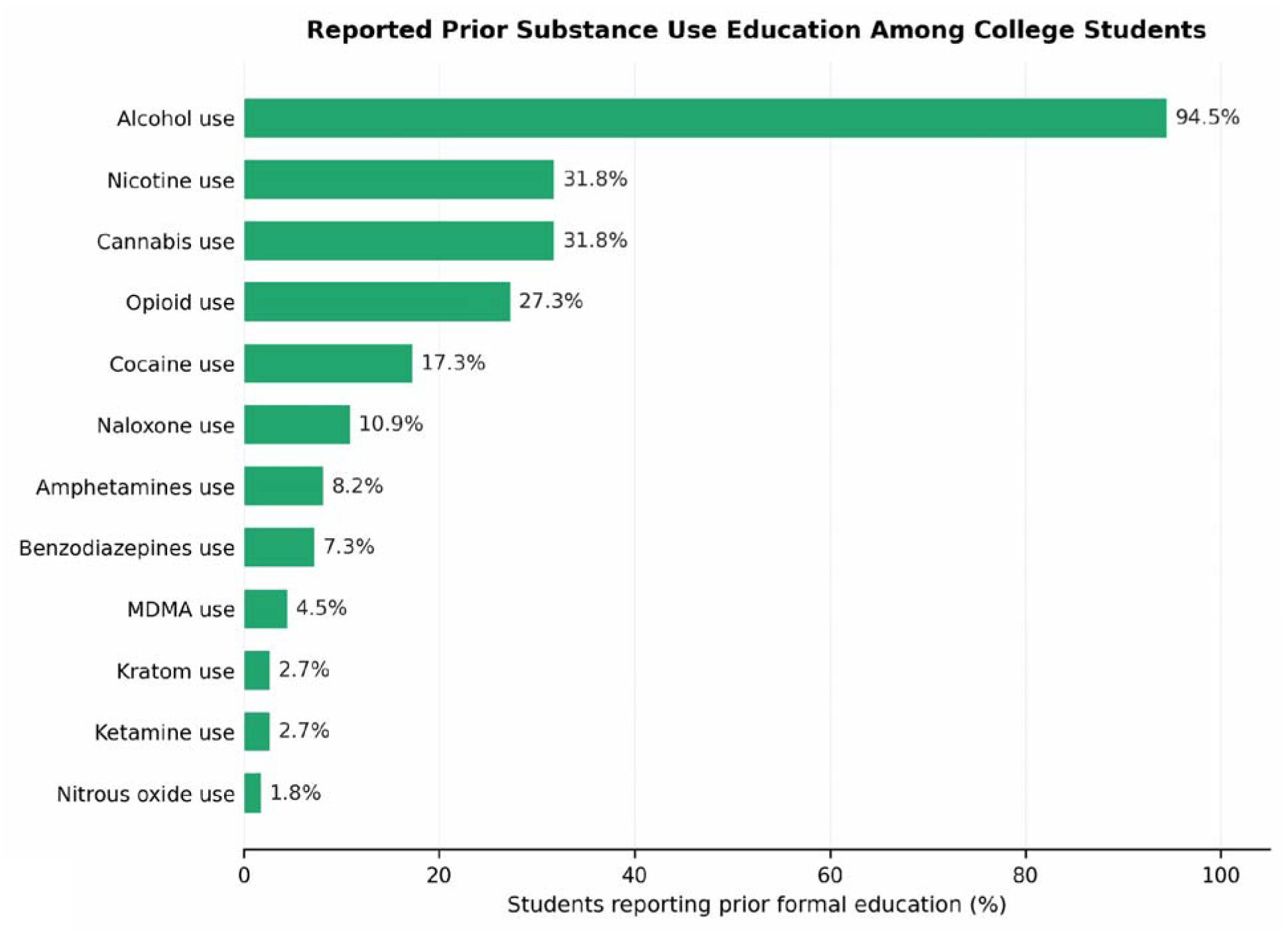
Prior formal substance education reported at pre-survey. Percentage of students reporting prior formal education on each of 12 substances/topics, among all date-eligible pre-survey respondents at the University of Nebraska-Lincoln and Team Awareness Combating Overdose (TACO) Inc. combined (n=110; pre-survey completed on or before March 31, 2026). Students selected all topics on which they had ever received formal education. Alcohol education was near-universal (94.5%); education on naloxone (10.9%), cocaine (17.3%), benzodiazepines (7.3%), and MDMA (4.5%) was uncommon.

**Figure 2.**
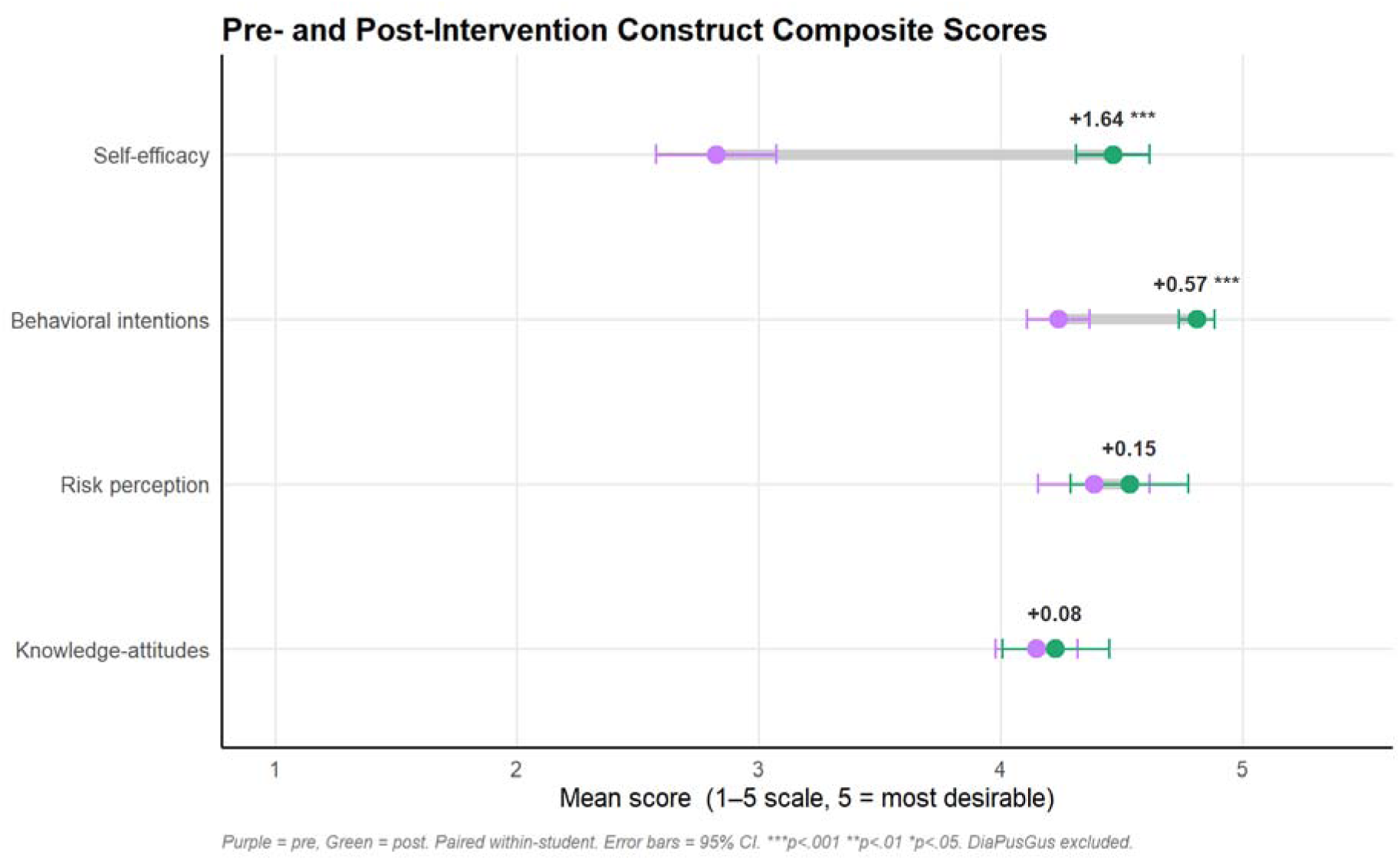
Construct composite scores before and immediately after Portal completion. Paired pre- and immediate post-intervention means for the four construct composites (5-point scales; N=42 paired respondents across both sites). Self-efficacy (7 items, d=2.00) and behavioral intentions (5 items, d=1.43) improved significantly (Benjamini-Hochberg adjusted p<.001, paired t-tests); risk perception and knowledge/attitudes began near the scale ceiling (4.38 and 4.15) and did not change significantly. Effect sizes are Cohen’s d for paired data.

**Figure 3.**
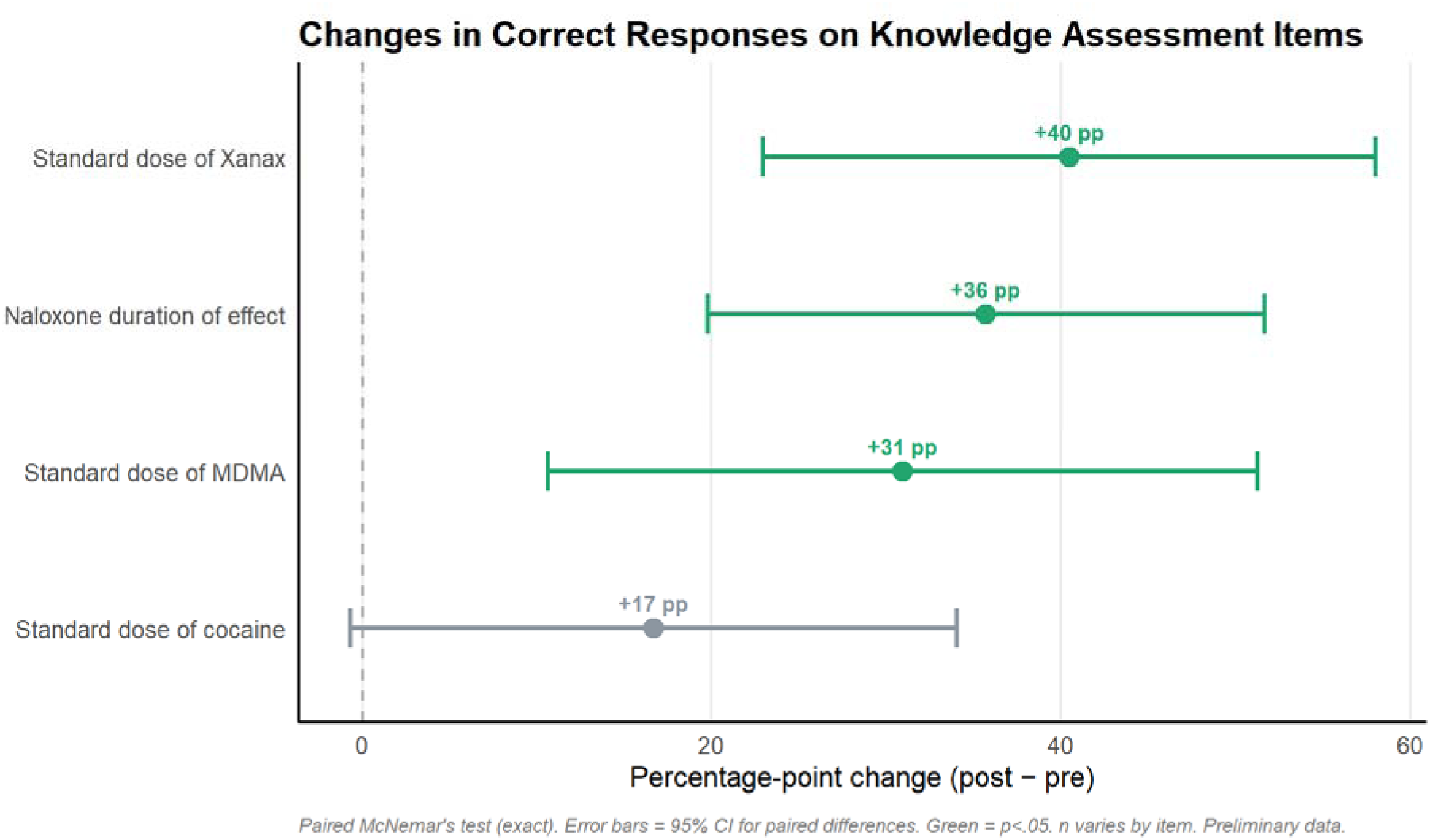
Change in factual knowledge accuracy from pre- to immediate post-intervention. Percentage-point change in the proportion of students answering each of four numeric-recall knowledge questions correctly (N=42; exact McNemar tests with Benjamini-Hochberg correction across all eight primary tests). Naloxone duration of action (+35.7pp), alprazolam standard dose (+40.5pp), and MDMA standard dose (+31.0pp) improved significantly (adjusted p=.017 or smaller); the cocaine standard dose item (+16.7pp) did not reach significance (adjusted p=.16).

**Figure 4.**
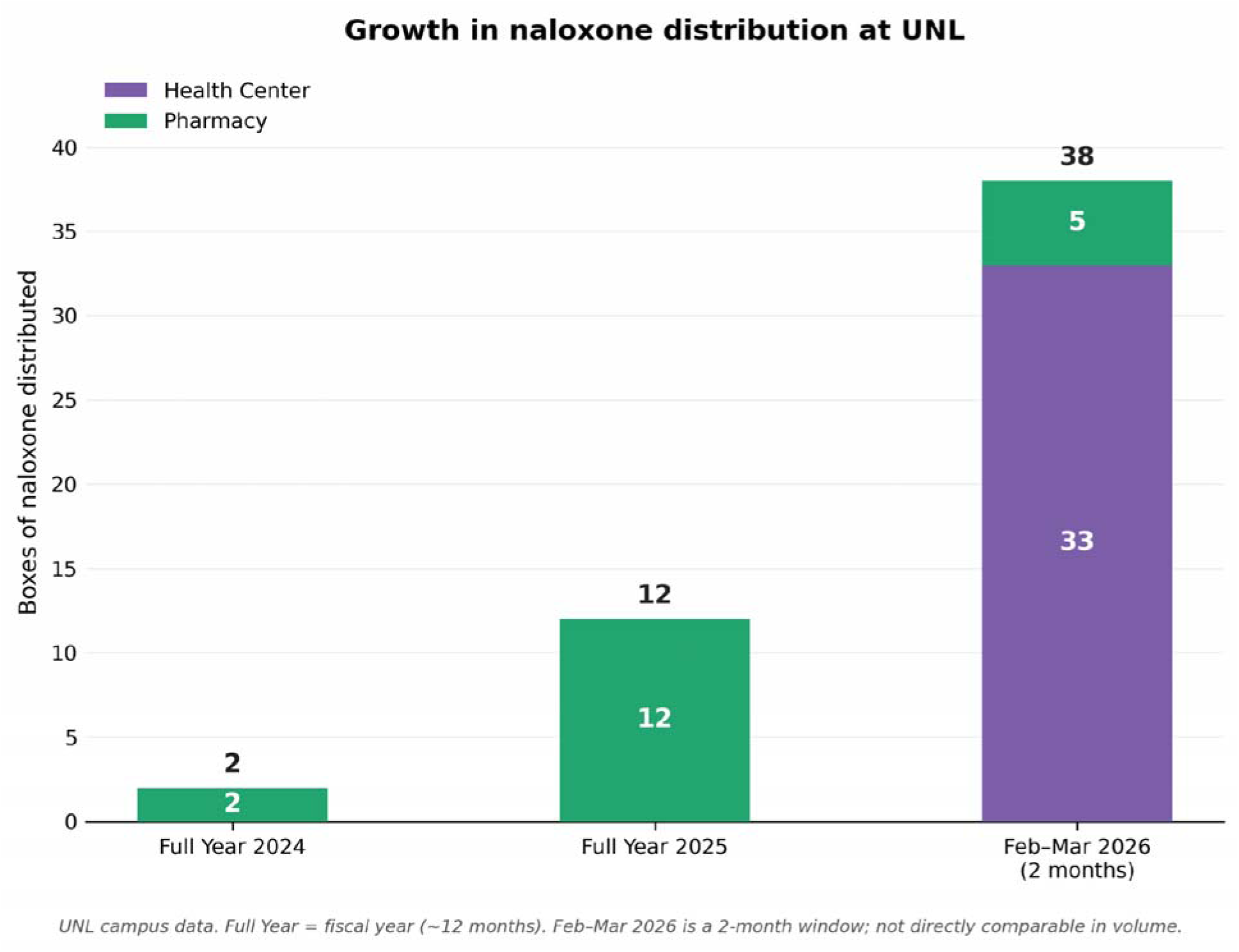
Naloxone kits distributed at the University of Nebraska-Lincoln, by period and source. Kits distributed by the campus pharmacy and the student health center: calendar year 2024 (2 kits, pharmacy only), calendar year 2025 (12 kits, pharmacy only; the health center stocked naloxone throughout 2025 and distributed none), and February 1–March 31, 2026, the first two months following Portal deployment (38 kits: 5 pharmacy, 33 health center). Distribution was limited to one kit per person; counts are institutional totals not linked to individual Portal completers.

### 3.6 Naloxone acquisition (ecological, UNL)

In the two months following Portal deployment, 38 naloxone kits were distributed, including 33 from a health center office that had stocked naloxone throughout 2025 and distributed none.

Distribution was capped at one kit per person, although there was no formal process to prevent students from accessing 1 kit from each location. No concurrent awareness campaigns, policy changes, or procedural changes occurred. The four in-person trainings of 2025 had produced no comparable acquisition behavior. The Portal includes no in-person component; 49 students completed it during this window. These data are institutional-level and not linked to individual completers; the count plausibly includes peers of completers, which is consistent with the peer-modeling mechanism discussed below. Overall, the number of naloxone kits distributed during the two-month period observed was more than double that of the prior 24 months combined.

**Table 5.** Naloxone kits distributed at UNL, by period and source.

| Period | Source | Kits distributed |
| --- | --- | --- |
| Calendar 2024 | Pharmacy only | 2 |
| Calendar 2025 | Pharmacy only (health center: 0) | 12 |
| Feb 1 – Mar 31, 2026 | Pharmacy (5) + health center (33) | 38 |

### 3.7 Acceptability

Among paired respondents, 95.2% (40/42) agreed or strongly agreed that Portal content would be useful in real life (UNL 25/27; TACO 15/15), and 93.3% (14/15 of those shown the later-added item) would recommend it. At the mandated high-school site, 100% (94/94) agreed or strongly agreed the Portal taught them something new about overdose prevention and 96.8% (91/94) that it was useful for their health. Across voluntary and mandated cohorts, no student gave a negative rating on any evaluation item.

## 4. DISCUSSION

### 4.1 Principal findings

A fully digital, gamified, mastery-based platform produced very large gains in substance-related self-efficacy (d=2.00; conservative sensitivity estimate d=1.60) and overdose-response behavioral intentions (d=1.43), and significant gains on three of four numeric-recall knowledge items, in a paired sample spanning two sites with distinct motivation profiles. Effects survived false-discovery correction and did not differ by site or by completion pacing. Concurrently, campus naloxone distribution at the primary site increased from zero health-center kits in all of 2025 to 38 total kits in two months – an ecological signal consistent with educational gains translating into protective behavior. Acceptability was uniform: across voluntary and mandated cohorts, not one student rated the Portal negatively.

### 4.2 DopaGE Portal yields change in self-efficacy

Students arrived already convinced that these substances are dangerous: baseline risk perception averaged 4.38 of 5 and did not change. At baseline, students’ capability for taking safety-promoting actions in situations related to substance use was insufficient. Following use of the Portal, average self-efficacy improved from 2.82 to 4.46. This pattern was maintained within the self-efficacy composite: the party-scenario refusal item began at ceiling (4.43) and stayed there, while substance-specific safety confidence and response confidence rose dramatically (d=1.18–2.80 across items). This dissociation is informative for prevention design. Programs built to convince young adults that drugs are dangerous or to rehearse refusal target constructs that, by these data, were already saturated. These data raise a question for future study: whether such baseline confidence may be inflated. Students expressing high refusal confidence and high risk awareness answered factual questions at 24–38% accuracy, a confidence-knowledge dissociation that the Portal improved from the knowledge side. Skills-based, informatively comprehensive education addressed the actual deficit.

In providing education to protect against potentially life-threatening scenarios to young adults, their cognitive developmental stage of late adolescence should not only inform what is taught, but also how it is presented. Directive prohibition failed its evaluations [14] in the population where autonomy is the central developmental task [21,22]. The Portal respects the autonomy students demonstrate, as refusal confidence was already near ceiling, while supporting the growth of previously limited capability: if a student does decide to experiment, information-complete education leaves them able to recognize contamination risk, avoid lethal combinations, and respond to an overdose, whereas prohibition-only education leaves them nothing to support survival of such a decision to use. Meanwhile, for students with some or no interest in substance experimentation, comprehensive education aims to provide them with specific evidence to determine their risk versus benefit assessment of substance use toward forming a more durable, autonomous decision not to use.

### 4.3 The behavioral signal and its mechanism

The naloxone finding deserves careful interpretation in both directions. It is ecological: kits were not linked to individuals, and we cannot attribute an acquisition to any particular participant within this study. But its timing, magnitude, and context are a strong signal: a health center that distributed zero kits over 12 months of stocked availability then distributed 33 in two months; the only identifiable new variable was a digital platform with no in-person component; the prior year’s in-person trainings had produced no equivalent behavior. Of 49 Portal completers, 38 acquisition events may imply reach beyond the participant population, consistent with self-efficacy theory’s peer-modeling mechanism that holds particular strength in childhood and adolescence [18], in which trained peers become visible models for protective behavior within their networks. Naloxone is rarely self-administered; acquiring it is predominantly a community-protective act. We frame this signal as hypothesis-generating: a future randomized trial will test individual-level behavioral translation directly.

This pattern mirrors the national one for young adults (Section 1.4): awareness and access without capability leave naloxone possession below 5% [12] and mortality unmoved [13]; here, access alone (2025) produced little to no action, while education produced acquisition.

Capability, not availability, appears to be the binding constraint.

### 4.4 Comparison with existing interventions

Three comparator educational mechanisms frame these results. In-person overdose education and naloxone distribution programs are effective: a meta-analysis found that trained bystanders successfully recognize and respond to overdoses [33], but they require trained instructors, a repeated process of mass scheduling, and physical presence – operational fragilities our primary site’s discontinued 2025 trainings illustrate. Classroom prevention curricula can produce knowledge gains – a 14-session teacher-delivered pilot improved overdose-detection knowledge from 1.0% to 41.7% [34] – but their authors identify teacher quality as the most influential variable, an inherent fidelity constraint. The Portal eliminates instructor variability entirely: delivery is identical for every student, invariant at scale, and as the site comparison shows, robust to how students pace themselves through to completion. Passive digital programs scale but underperform: AlcoholEdu’s randomized evaluations show small knowledge effects and limited positive behavioral effects not demonstrated beyond 30 days [15,16]. Against this grid, the Portal’s combination of digital scale, mastery-based active learning, substance-comprehensive content, and autonomy-respecting framing [35] occupies a previously untested cell, and these preliminary results suggest this cell is worth further testing with additional rigor.

### 4.5 Strengths and limitations

Strengths include the paired design with anonymous self-generated identification code (SGIC) linkage; replication across two independently motivated populations from the same age cohort with formal poolability testing; an ecological behavioral outcome that extends inference beyond self-report; false-discovery correction across all primary tests; a pre-specified sensitivity composite confirming robustness; instruments adapted from validated scales [27–32]; an independent reanalysis reproducing all values from raw data; and acceptability evidence from a mandated cohort free of self-selection.

Limitations are substantial and define future studies. The design is single-group pre-post: maturation, testing effects, and demand characteristics cannot be excluded, and causal language is unwarranted. The paired sample is modest (N=42). No demographic data were collected, precluding subgroup analysis. The naloxone outcome is institutional-level, not individual-linked. TACO as a supporting site exclusively contains students newly self-selected into a substance-use-prevention volunteer role within the past 2 months and therefore carries a different motivation profile than a general student body. A certificate-delivery defect (fixed March 26, 2026) meant pre-fix iPost completion was 57.4% at UNL and 55.6% at TACO versus 100% post-fix, so pre-fix data may over-represent more persistent students. The survey item on potential for recommendation to others was added mid-study (lower n). Ninety-day follow-up was insufficient for analysis (n=6), though ongoing collection suggests longitudinal assessment is feasible on this platform. The high-school cohort used a similar length but overall different, school-administered instrument, limiting comparability to three items that remained identical to those in our instrument. Self-selection into a voluntary platform limits generalizability of the college results, though partially offset by the uniformly positive mandated-cohort ratings.

### 4.6 Implications and next steps

The epidemiology argues that the decisive variable in young-adult overdose mortality is shifting toward the scene of the event: fentanyl involvement is near 4 in 5 [1], most fatal events presenting missed intervention opportunities [5], and non-fatal events at record highs [2]. The education system serving this population teaches almost none of the most relevant capability, and the dominant digital model is passive and ignores illicit substance use where fentanyl involvement resides [15–17]. These data show that a mastery-based digital alternative has the potential to produce much-needed capability improvements, at a delivery cost and fidelity that in-person programs cannot match, with a behavioral signal that access-alone policies have not produced. Regional variation in contamination [7] further argues that definitive evaluation must be multi-site and regionally diverse. Such evaluation is now intended: a multi-site randomized controlled trial with an attention-matched and non-gamified control, longitudinal follow up at both 90- and 180-days, and individual-level behavioral outcomes.

## 5. CONCLUSION

Young adults (ages 18-25) have little need to be persuaded that substance use is dangerous; they need to be equipped to act in preventing such dangers. In this preliminary evaluation, a gamified, mastery-based, fully digital platform produced large, site-consistent gains in overdose-response self-efficacy and substance-specific knowledge, left already-saturated risk perception consistent, and was followed immediately by a campus-level surge in naloxone acquisition that stocked availability with or without in-person naloxone trainings had never produced. These findings, and their specific limitations, justify a future randomized controlled trial.

## DECLARATIONS

### Funding

Deployment of the DopaGE Portal at the University of Nebraska-Lincoln is funded by an Obvious Expenditure Grant from the Nebraska Region V Systems Opioid Abatement Funds awarded to DopaGE Inc. for the purpose of providing the training to first-year and transfer students in fiscal year 2026. Grant outcome measures, proposed by DopaGE and adopted by the funder, comprise Portal completion rates, immediate post- and 90-day survey completion rates, and campus naloxone distribution counts; survey responses and results are not grant deliverables. The funder had no access to survey data and no role in study design, analysis, or manuscript preparation. Study design, analysis, and writing were funded by DopaGE.

### Competing interests

M.H. is the founder and CEO of DopaGE Inc., which develops the Portal, and the founder and Executive Chairwoman of Team Awareness Combating Overdose (TACO) Inc., an independent 501(c)(3) non-profit with no ownership relationship to DopaGE; TACO purchased Portal access from DopaGE using donation revenue. R.F. serves as a Board Advisor to TACO. R.F., S.H., E.Z., and T.K. hold stock options in DopaGE under vesting agreements. A.M. is TACO’s Director of Grants and a DopaGE intern; she completed the Portal as a TACO volunteer prior to the study window, and no author’s data appear in the study sample.

### Ethics

See Methods 2.8. The project was reviewed by the University of Nebraska-Lincoln IRB office and determined not to require full review; data are de-identified under 45 CFR 46.104(d)(4).

## Data Availability

De-identified response-level data and analysis code are available from the corresponding author on reasonable request.

https://wonder.cdc.gov/controller/saved/D176/D509F527

https://wonder.cdc.gov/controller/saved/D176/D509F526

https://wonder.cdc.gov/controller/saved/D176/D509F532

https://wonder.cdc.gov/controller/saved/D176/D509F537

https://wonder.cdc.gov/controller/saved/D176/D509F534

https://wonder.cdc.gov/controller/saved/D176/D509F679

https://wonder.cdc.gov/controller/saved/D176/D509F560

https://wonder.cdc.gov/controller/saved/D176/D509F653

https://nemsis.org/drug-overdose-surveillance-dashboard/

https://www.cdc.gov/overdose-prevention/data-research/facts-stats/sudors-dashboard-fatal-overdose-data.html

## Acknowledgments

The authors thank Anusha Puri for background research and comments on the manuscript.

## Authors’ contributions

**Conceptualization:** M.H., R.F., T.K.

**Platform design and development:** M.H., with clinical oversight from S.H. and E.Z. and contributing background research from R.F.

**Survey instrument development:** M.H., R.F., T.K.

**Statistical design:** R.F.

**Analysis code and figures:** A.M., supervised by R.F.

**Clinical content validation:** E.Z., S.H.

**Drafting:** M.H.

**Review and editing:** all authors.

**Project administration:** M.H.

**Supervision:** S.H.

All authors reviewed and approved the final manuscript.

## SUPPLEMENTARY MATERIALS

**Table S1.** Individual Likert item results (all 24 items, pre/post means, d).

| Item | Pre M | Post M | d | p (BH) |
| --- | --- | --- | --- | --- |
| **Self-Efficacy (7 items)** |  |  |  |  |
| Would avoid drug use in party scenario | 4.43 | 4.36 | -0.06 | .82 |
| Confident checking overdose signs | 3.21 | 4.4 | +1.00 | <.001*** |
| Confident administering naloxone | 3.07 | 4.64 | +1.24 | <.001*** |
| Confident using cocaine dosing knowledge for safety | 2.93 | 4.5 | +1.18 | <.001*** |
| Confident using MDMA dosing knowledge for safety | 2.26 | 4.38 | +1.41 | <.001*** |
| Well-educated to help friend stay safe using Xanax | 1.9 | 4.52 | +2.80 | <.001*** |
| Well-educated to help friend stay safe using MDMA | 1.95 | 4.43 | +2.01 | <.001*** |
| **Behavioral Intentions (5 items)** |  |  |  |  |
| Would call ambulance right away | 4.4 | 4.95 | +0.92 | <.001*** |
| Wouldn't take further action beyond calling [R] | 3.76 | 4.6 | +0.98 | <.001*** |
| Would stay with victim until help arrives | 4.52 | 4.86 | +0.49 | .006** |
| Understand utility of Good Samaritan Laws | 3.98 | 4.81 | +0.74 | <.001*** |
| Would feel safer if peers trained in overdose response | 4.52 | 4.83 | +0.60 | <.001*** |
| **Risk Perception (6 items)** |  |  |  |  |
| Risk to health trying cocaine once or twice | 4.21 | 4.29 | +0.05 | .82 |
| Risk to health using cocaine regularly | 4.74 | 4.83 | +0.22 | .24 |
| Risk to health trying Xanax without prescription | 4.05 | 4.17 | +0.10 | .69 |
| Risk to health using Xanax without prescription regularly | 4.5 | 4.79 | +0.39 | .03* |
| Risk to health trying MDMA once or twice | 4.24 | 4.29 | +0.04 | .83 |
| Risk to health using MDMA regularly | 4.57 | 4.83 | +0.53 | .003** |
| **Knowledge/Attitudes (6 items)** |  |  |  |  |
| Alcohol + cocaine combination is riskier [R] | 4.02 | 3.9 | -0.06 | .82 |
| Cocaine contamination with other substances is common [R] | 4.48 | 4.9 | +0.58 | .001** |
| Easy to become dependent on Xanax | 4.4 | 4.64 | +0.30 | .09 |
| Alcohol + Xanax combination is riskier [R] | 4.4 | 4.24 | -0.10 | .69 |
| Alcohol + MDMA combination is riskier [R] | 3.93 | 4.02 | +0.05 | .82 |
| Would decline offered prescription drug from friend | 3.64 | 3.64 | +0.00 | 1.0 |

[R] = reverse-keyed item. d = Cohen's d for paired data. p (BH) = Benjamini-Hochberg adjusted p-value across all 24 items. \*\*\*p<.001 \*\*p<.01 \*p<.05.

**Table S2.** Hockaday high-school cohort: pre- and post-portal Likert item results and post-completion assessment (Spring 2026; PRE n=98, POST n=94).

| Item | Pre M<br>[n=98] | Post M<br>[n=94] | Post<br>%A+SA |
| --- | --- | --- | --- |
| **Knowledge, Confidence & Behavioral Intention (pre/post paired; n=98 pre, n=94 post)** |  |  |  |
| I would be able to identify if someone was experiencing a drug overdose. | 3.18 | 4.13 | 96.8% |
| I am confident in my ability to help someone experiencing a drug overdose. | 4.02 | 3.99 | 87.2% |
| I know how to administer naloxone (Narcan) to someone experiencing a drug overdose. | 1.91 | 4.41 | 98.9% |
| I feel confident in my ability to reduce my risk of harm in a situation involving substance use. | 3.40 | 4.32 | 97.9% |
| If offered a substance in a social setting, I feel confident in my ability to say no. | 4.66 | 4.73 | 98.9% |
| **Post-Portal Assessment (post-only; n=94)** |  |  |  |
| This portal taught me something about overdose prevention that I did not already know. | — | 4.81 | 100% |
| This portal is something that I found useful for my health and/or for the health of those around me. | — | 4.72 | 96.8% |

Items use 5-point Likert scale (1 = Strongly Disagree to 5 = Strongly Agree). PRE n=98; POST n=94. Post-only items were administered immediately following portal completion and have no pre-survey comparison. %A+SA = percentage endorsing Agree or Strongly Agree.

**Table S3.**
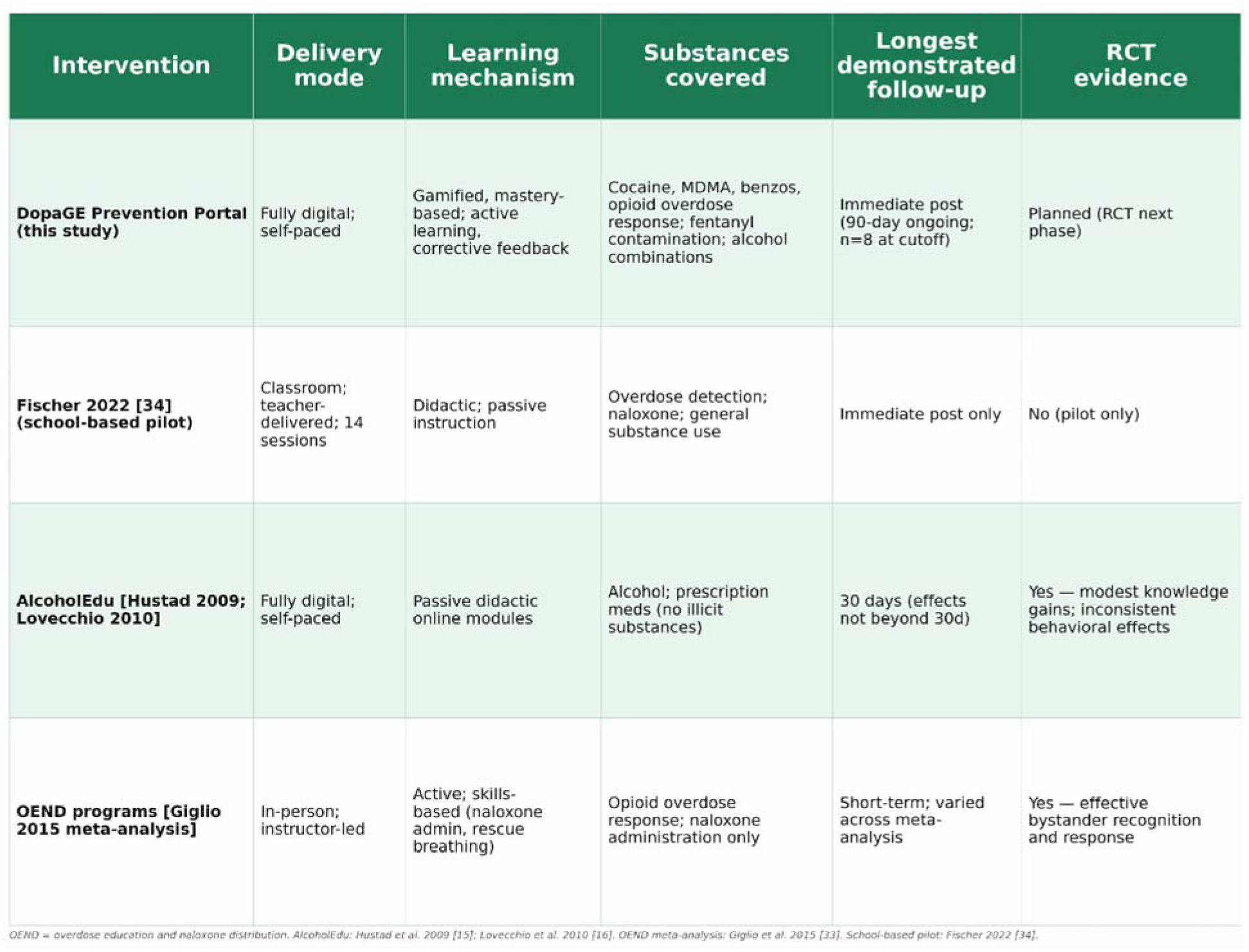
Comparable interventions grid (delivery mode × mechanism × substances covered × longest demonstrated follow-up): this study; Fischer 2022 [34]; AlcoholEdu trials [15,16]; OEND programs [33].

| Intervention | Delivery mode | Learning mechanism | Substances covered | Longest demonstrated follow-up | RCT evidence |
| --- | --- | --- | --- | --- | --- |
| **DopaGE Prevention Portal (this study)** | Fully digital; self-paced | Gamified, mastery-based; active learning, corrective feedback | Cocaine, MDMA, benzos, opioid overdose response; fentanyl contamination; alcohol combinations | Immediate post (90-day ongoing; n=8 at cutoff) | Planned (RCT next phase) |
| **Fischer 2022 [34] (school-based pilot)** | Classroom; teacher-delivered; 14 sessions | Didactic; passive instruction | Overdose detection; naloxone; general substance use | Immediate post only | No (pilot only) |
| **AlcoholEdu [Hustad 2009; Lovecchio 2010]** | Fully digital; self-paced | Passive didactic online modules | Alcohol; prescription meds (no illicit substances) | 30 days (effects not beyond 30d) | Yes — modest knowledge gains; inconsistent behavioral effects |
| **OEND programs [Giglio 2015 meta-analysis]** | In-person; instructor-led | Active; skills-based (naloxone admin, rescue breathing) | Opioid overdose response; naloxone administration only | Short-term; varied across meta-analysis | Yes — effective bystander recognition and response |

OEND = overdose education and naloxone distribution. AlcoholEdu: Hustad et al. 2009 [15]; Lovecchio et al. 2010 [16]. OEND meta-analysis: Giglio et al. 2015 [33]. School-based pilot: Fischer 2022 [34].

**Table S4.**
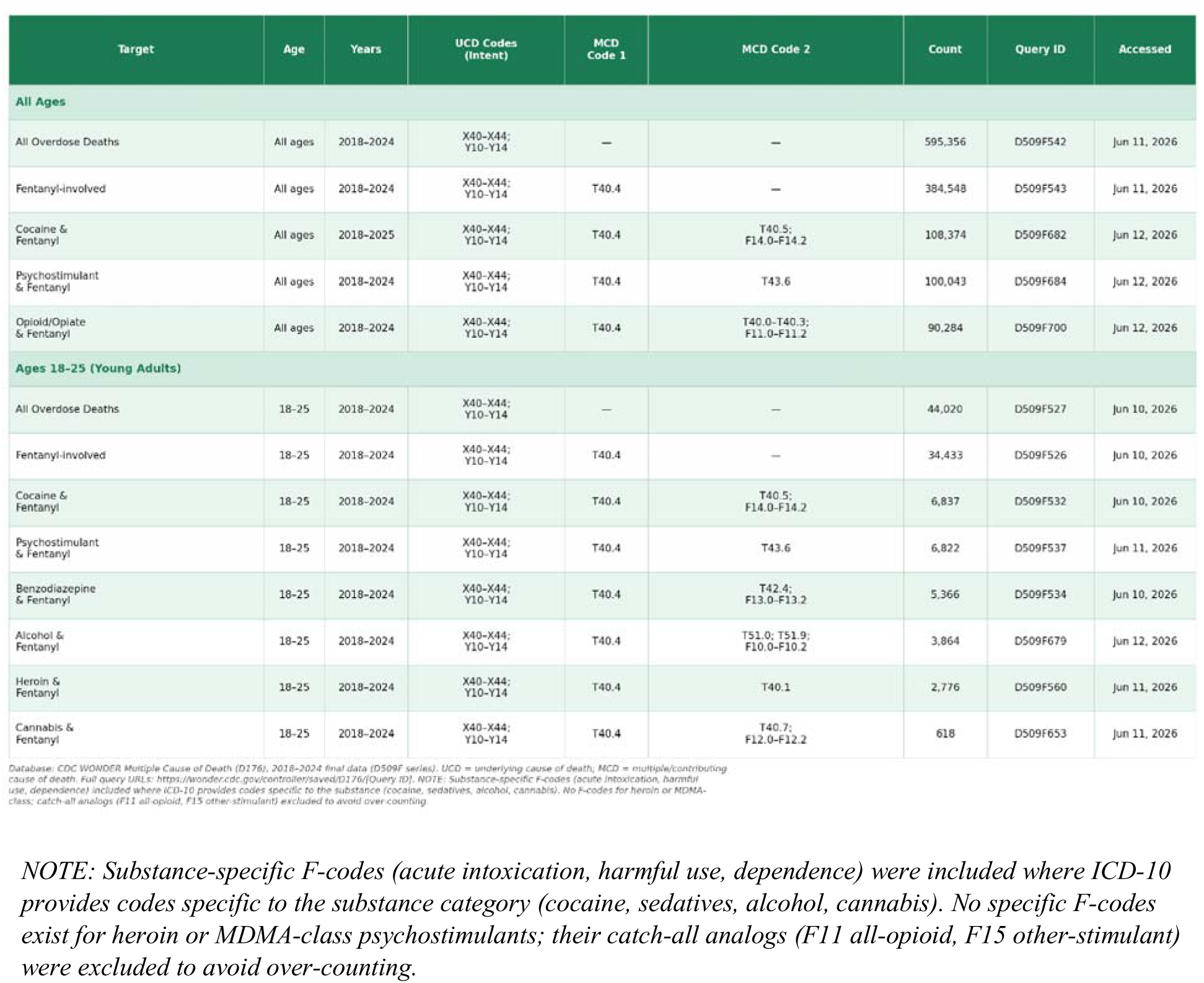
CDC WONDER query specifications: database, years, age range, intent codes (UCD), multiple-cause ICD-10 codes per substance category, counts, and saved-query URLs for all mortality statistics cited in the Introduction.

| Target | Age | Years | UCD Codes (Intent) | MCD Code 1 | MCD Code 2 | Count | Query ID | Accessed |
| --- | --- | --- | --- | --- | --- | --- | --- | --- |
| **All Ages** |  |  |  |  |  |  |  |  |
| All Overdose Deaths | All ages | 2018–2024 | X40–X44;<br>Y10–Y14 | — | — | 595,356 | D509F542 | Jun 11, 2026 |
| Fentanyl-involved | All ages | 2018–2024 | X40–X44;<br>Y10–Y14 | T40.4 | — | 384,548 | D509F543 | Jun 11, 2026 |
| Cocaine & Fentanyl | All ages | 2018–2025 | X40–X44;<br>Y10–Y14 | T40.4 | T40.5;<br>F14.0–F14.2 | 108,374 | D509F682 | Jun 12, 2026 |
| Psychostimulant & Fentanyl | All ages | 2018–2024 | X40–X44;<br>Y10–Y14 | T40.4 | T43.6 | 100,043 | D509F684 | Jun 12, 2026 |
| Opioid/Opiate & Fentanyl | All ages | 2018–2024 | X40–X44;<br>Y10–Y14 | T40.4 | T40.0–T40.3;<br>F11.0–F11.2 | 90,284 | D509F700 | Jun 12, 2026 |
| **Ages 18–25 (Young Adults)** |  |  |  |  |  |  |  |  |
| All Overdose Deaths | 18–25 | 2018–2024 | X40–X44;<br>Y10–Y14 | — | — | 44,020 | D509F527 | Jun 10, 2026 |
| Fentanyl-involved | 18–25 | 2018–2024 | X40–X44;<br>Y10–Y14 | T40.4 | — | 34,433 | D509F526 | Jun 10, 2026 |
| Cocaine & Fentanyl | 18–25 | 2018–2024 | X40–X44;<br>Y10–Y14 | T40.4 | T40.5;<br>F14.0–F14.2 | 6,837 | D509F532 | Jun 10, 2026 |
| Psychostimulant & Fentanyl | 18–25 | 2018–2024 | X40–X44;<br>Y10–Y14 | T40.4 | T43.6 | 6,822 | D509F537 | Jun 11, 2026 |
| Benzodiazepine & Fentanyl | 18–25 | 2018–2024 | X40–X44;<br>Y10–Y14 | T40.4 | T42.4;<br>F13.0–F13.2 | 5,366 | D509F534 | Jun 10, 2026 |
| Alcohol & Fentanyl | 18–25 | 2018–2024 | X40–X44;<br>Y10–Y14 | T40.4 | T51.0; T51.9;<br>F10.0–F10.2 | 3,864 | D509F679 | Jun 12, 2026 |
| Heroin & Fentanyl | 18–25 | 2018–2024 | X40–X44;<br>Y10–Y14 | T40.4 | T40.1 | 2,776 | D509F560 | Jun 11, 2026 |
| Cannabis & Fentanyl | 18–25 | 2018–2024 | X40–X44;<br>Y10–Y14 | T40.4 | T40.7;<br>F12.0–F12.2 | 618 | D509F653 | Jun 11, 2026 |

Database: CDC WONDER Multiple Cause of Death (D176); 2018–2024 final data (D509F series). UCD = underlying cause of death; MCD = multiple/contributing cause of death. Full query URLs: https://wonder.cdc.gov/controllers/saved/D176/QueryID. NOTE: Substance-specific F-codes (acute intoxication, harmful use, dependence) included where ICD-10 provides codes specific to the substance (cocaine, sedatives, alcohol, cannabis). No F-codes for heroin or MDMA-class; catch-all analogs (F11 all-opioid, F15 other-stimulant) excluded to avoid over-counting.
**NOTE:** Substance-specific F-codes (acute intoxication, harmful use, dependence) were included where ICD-10 provides codes specific to the substance category (cocaine, sedatives, alcohol, cannabis). No specific F-codes exist for heroin or MDMA-class psychostimulants; their catch-all analogs (F11 all-opioid, F15 other-stimulant) were excluded to avoid over-counting.

## Notes

### Author Declarations

IRB of The University of Nebraska-Lincoln waived ethical approval for this work.

